# Organising outpatient dialysis services during the COVID-19 pandemic. A simulation and mathematical modelling study

**DOI:** 10.1101/2020.04.22.20075457

**Authors:** Michael Allen, Amir Bhanji, Jonas Willemsen, Steven Dudfield, Stuart Logan, Thomas Monks

**Affiliations:** University of Exeter Medical School & NIHR South West Peninsula Applied Research Collaboration (ARC); Portsmouth Hospitals, NHS Trust; University of Exeter Medical School

## Abstract

**Background:** This study presents two simulation modelling tools to support the organisation of networks of dialysis services during the COVID-19 pandemic. These tools were developed to support renal services in the South of England (the Wessex region caring for 650 patients), but are applicable elsewhere.

**Methods:** A discrete-event simulation was used to model a worst case spread of COVID-19 (80% infected over three months), to stress-test plans for dialysis provision throughout the COVID-19 outbreak. We investigated the ability of the system to manage the mix of COVID-19 positive and negative patients, and examined the likely effects on patients, outpatient workloads across all units, and inpatient workload at the centralised COVID-positive inpatient unit. A second Monte-Carlo vehicle routing model estimated the feasibility of patient transport plans and relaxing the current policy of single COVID-19 patient transport to allow up to four infected patients at a time.

**Results:** If current outpatient capacity is maintained there is sufficient capacity in the South of England to keep COVID-19 negative/recovered and positive patients in separate sessions, but rapid reallocation of patients may be needed (as sessions are cleared of negative/recovered patients to enable that session to be dedicated to positive patients). Outpatient COVID-19 cases will spillover to a secondary site while other sites will experience a reduction in workload. The primary site chosen to manage infected patients will experience a significant increase in outpatients and in-patients. At the peak of infection, it is predicted there will be up to 140 COVID-19 positive patients with 40 to 90 of these as inpatients, likely breaching current inpatient capacity (and possibly leading to a need for temporary movement of dialysis equipment).

Patient transport services will also come under considerable pressure. If patient transport operates on a policy of one positive patient at a time, and two-way transport is needed, a likely scenario estimates 80 ambulance drive time hours per day (not including fixed drop-off and ambulance cleaning times). Relaxing policies on individual patient transport to 2-4 patients per trip can save 40-60% of drive time. In mixed urban/rural geographies steps may need to be taken to temporarily accommodate renal COVID-19 positive patients closer to treatment facilities.

**Conclusions:** Discrete-event simulation simulation and Monte-Carlo vehicle routing model provides a useful method for stress-testing inpatient and outpatient clinical systems prior to peak COVID-19 workloads.

## 1 Introduction

Severe Acute Respiratory Syndrome-Corona Virus-2 (SARS-CoV-2) COVID-19 (henceforth known as COVID) is causing widespread disruption to normal healthcare services, as the number COVID-positive cases increases. In the UK a worst case scenario is that 80% of the population are infected over a three month period, if controls are not put in place [1]. Although social distancing measures are in place both in the UK and internationally, patients with Chronic Kidney Disease who must visit dialysis units are limited in their ability to be fully isolated. It is possible therefore that spread in the dialysis population will be faster than in the general population.

Rapid guidelines for dialysis service delivery have been published[2, 3, 4]. These include separation of COVID-positive and COVID-negative patients; dialysis units working with transport providers to minimise the risk of cross-infection; and continuing to treat patients as close to home as possible. [2].

Planning service delivery that separates COVID-positive patients is complicated, due to the uncertainty of the spread of SARS-CoV-2, the variability seen in symptom onset, length of infectivity, and regional delivery of dialysis.

We therefore sought to support decision making in the period prior to peak infection by developing mathematical models of dialysis service delivery and patient transport. We aimed to provide reusable tools to provide rapid information under various scenarios including a worst case three month spread.

## 2 Methods

We developed a discrete-event simulation (DES) model of service delivery in the dialysis network. DES is an appropriate method to capture the stochastic dynamics of a capacity constrained system and model patients individually [5]. DES has been applied extensively in health service delivery [6, 7, 8, 9] and previously to model dialysis demand [10] as well as networks of care facilities [11]. We also developed a Monte-Carlo vehicle routing model to model patient transport. The algorithm used, a combination of the Clarke-Wright Savings [12] method and Iterated Local Search [13], finds good solutions grouping and ordering patient pickup.

### 2.1 Study Setting

We apply the service delivery modelling tools [14] in the South of England in the region of Wessex: a mixed urban/rural setting where the renal dialysis service cares for 644 patients. The service operates a network of nine centres. The largest of which is located at the Queen Alexandra (QA) Hospital, Portsmouth. To access dialysis services 75% of patients make use of patient transport services. During the epidemic, COVID-positive patients will be treated separately from negative and recovered. The Queen Alexandra will be used as the primary site for positive outpatients and inpatients with spillover to a second site (Basingstoke) when capacity is insufficient. Patient transport services will provide COVID only ambulances with a policy of single patient transport.

In the analysis we excluded home patients (n = 80) and due to its separation from the mainland the Isle of Wight (n = 44).

The geography of units and patients is described in more detail in appendix A.

### 2.2 Outcome measures

We estimated the the change in outpatient and inpatient workload during the epidemic in terms of COVID-positive negative and recovered, at each dialysis unit in the network. Estimates were produced over periods three to six months. We also estimated the number of patients who were required to travel to a different unit from normal and the change in travel time.

We estimated the vehicle total travel implications for patient transport services given a range of COVID-positive scenarios across the regions geography.

### 2.3 Data Sources

To ensure confidentiality, patient geographic locations was provided at the UK postcode sector level (alternatives might be output areas or northings and eastings). Travel times between these sectors were estimated using Routino (routino.org) with data from OpenStreetMap (openstreetmap.org).

The worst case time of spread of COVID-positive was taken from Fergeson et al. [1]. Mortality rate, time a patient was COVID-positive before admission and inpatient length of stay were local parameters.

### 2.4 Analysis environment

All models were written in Python 3.8. We used SimPy 3 [15] to implement the DES model. The transport model was implemented using pandas [16] and NumPy [17]. All charts were produced with MatPlotLib [18]. We provide all code and data used in the study and follow the STRESS reporting guidelines for DES [19]. The dialysis model results were run on an Intel i9-7980XE CPU with 64GB RAM running Ubuntu 19.10 Linux. The transport modelling results were run on an Intel i9-9900K CPU with 64GB RAM running the Pop! OS 19.10 Linux.

### 2.5 Verification and validation

We performed model testing (verification) as models were developed in line with simulation standards [20]. Two of the authors are experienced modellers and verification included a code review and cross working on models. Quantitative validation of models (checking models are appropriate detailed and sufficiently accurate) is challenging in the COVID epidemic as the forecast is of unprecedented conditions. We instead worked closely with clinicians, managers and informatics specialists within the local health system to review iterative versions of the model. We also opted to model a range of likely scenarios including what is widely believed to be the worst case.

### 2.6 Dialysis model

The dialysis model runs through a defined period (e.g. one year) and simulates the progression of patients through phases of COVID infection: negative, positive (with some requiring inpatient care) and recovered or died. The speed of progression of infection through the population may be varied (typically 3-12 months).

As patients change COVID state the model seeks to place them in the appropriate unit and session, opening up COVID-positive sessions in units that allow it. COVID-positive patients do not mix with any other patients. Opening up COVID-positive sessions causes other patients to be displaced from that session, and the model seeks to reallocate them either to the same unit or, if there is no space left, to the closest alternative unit.

The dialysis model is run 30 times to simulate 30 alternative years as, due to the randomness of infection, no two years will be exactly alike. Results show typical (median) and extreme years.

#### 2.6.1 Patient progression model

A simplification used in this model is that all patients should receive dialysis three times weekly, with each patient allocated to a starting day for the week of either Monday or Tuesday.

A proportion of patients moves through phases of COVID state and care (figure 1). The proportions of patients and times in each phase is either fixed or sampled from stochastic distributions as given in table 1. We assume that COVID patients must be separated from uninfected patients, and that patients who have recovered from a COVID episode do not mix with those currently testing COVID positive. We do not deal specifically with *suspected* COVID patients in the model, anticipating that rapid testing will soon be available to diagnose which group they belong to.

**Table 1:** Baseline model parameters

| Parameter | Distribution | Baseline values |
| --- | --- | --- |
| Proportion patients infected | Fixed | 0.8 |
| Time to infection | Normal | Mean = 60, SD = 15 (3 month spread) |
| Time positive/symptomatic (outpatient) | Uniform | Low = 7, High = 14 |
| Proportion COV requiring inpatient care | Fixed | 0.6 |
| Time inpatient COV before admission | Uniform | Low = 3, High = 7 |
| Time inpatient COV | Uniform | Low = 7, High = 14 |
| Mortality rate | Fixed | 0.15 |

**Figure 1:**
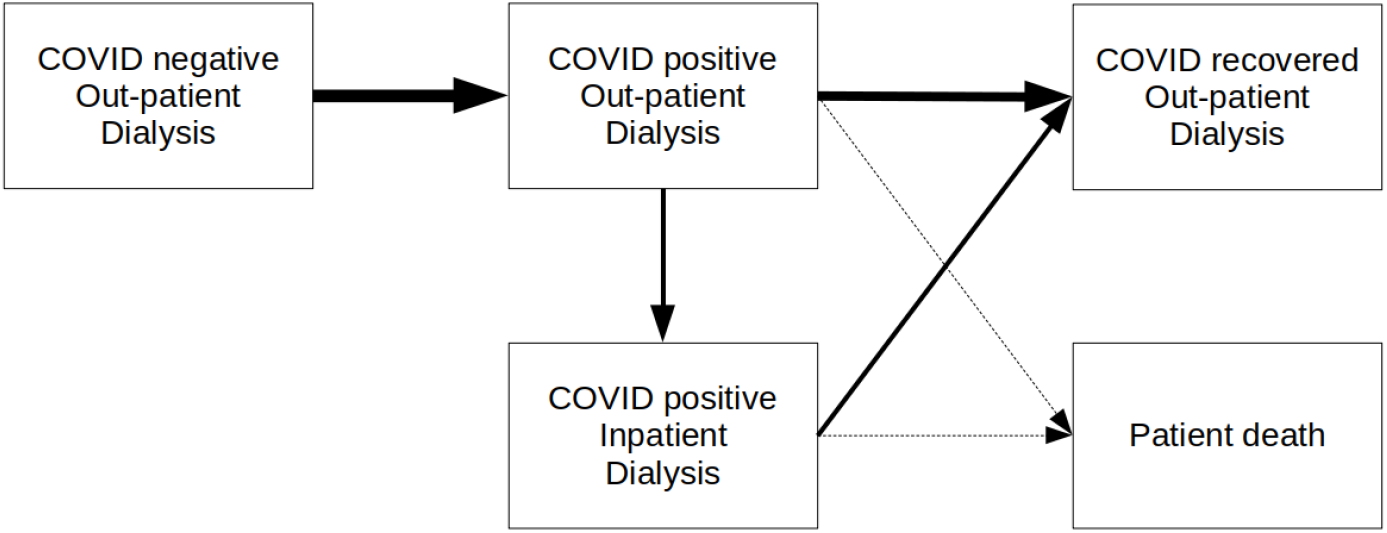
Schematic representation of patient pathway

The *baseline* model takes a *worst case* progression of COVID, infecting 80% of the dialysis population over 3 months.

#### 2.6.2 Unit search strategy

When allocating patients to units, the following search strategy is employed.

- *COVID negative*: First look for place in current unit attended. If no room there place in the closest unit (judged by estimated travel time) with available space.
- *COVID-positive*: Place all COVID-positive patients first in Queen Alexandra Hospital, Portsmouth, and if capacity there is fully utilised open up capacity in Basingstoke. If a new COVID session is required, the model will displace all COVID negative patients in that session, and seek to re-allocate them according to the rules for allocating COVID negative patients.
- *COVID-positive inpatient* : All inpatients are placed in Queen Alexandra Hospital, Portsmouth (though the model allows searching by travel time if another unit were to open to renal COVID-positive inpatients).
- *COVID-recovered* : Treat as COVID negative.
- *Unallocated patients*: If a patient cannot be allocated to any unit, the model attempts to allocate them each day.

Patients, in the model, may end up being cared for at a more distant unit than their starting unit. Once every week, the model seeks to reallocate patients back to their starting unit, or closest available unit if room in their starting unit is not available. This will also compress COVID-positive patients into as few units and sessions as possible.

COVID-positive sessions are converted back to COVID negative sessions when they are no longer needed.

### 2.7 Patient transport model

The transport model provides an estimate of the vehicle travel time needed to transport COVID-positive patients to (and from) an outpatient treatment facility.

#### 2.7.1 Transport scenarios

The model can vary the capacity of transport vehicles (e.g. the number of seats per ambulance) and the number of COVID positive patients in the population who need COVID-positive capable transport at any time.

We model the following daily scenarios:

- A population of 20 patients are COVID positive.
- A population of 40 patients are COVID positive.
- A population of 60 patients are COVID positive.
- Ambulances are able to pick up between 1 and 4 COVID positive patients on a single trip.

For example, if COVID spreads through the population in three months, there may be 140 COVID-positive patients. If 40% are inpatients, and 75% require transport, there may be 60-70 patients requiring COVID-positive transport (30-35 on each day).

#### 2.7.2 Simulation of pickup locations

There is no robust way to estimate which patients will become COVID positive and at what time. The model therefore uses a Monte-Carlo sampling approach to simulate different groups of patients becoming infected. The sampling uses the geographic distribution of patient home postcode sectors.

- We assume that all patients are equally likely to become infected.
- We weight the sample by the number of patients within each postcode sector. I.e. areas with more patients are more likely to be sampled.

The model works by performing multiple runs. On each run a different cohort of patients is selected. This means that hundreds of combinations of COVID-positive patient locations can be explored. The more common combinations will be sampled more frequently due to the weighting. For each sample a set of transport routes are created. The transport routes group patients together and order them for transport to the hospital. The grouping is based on travel time.

We simplified the problem to consider a symmetric road network. That is travel time outward to a patient is the same as inward travel time. In reality road networks are asymmetric, for example due to one way systems, and roadworks.

#### 2.7.3 Transport route construction

After a set of patient locations is chosen a set of routes are constructed. The number of routes needed depends on capacity of the ambulance. Each route has a home base for the ambulance. We have simplified the problem so that the ambulance is based at the QA (in reality it will start from an depot elsewhere, but this is only one leg of multiple journeys). In each scenario, we also simplified the problem so that all ambulances have the same capacity (no. of seats)

When vehicle capacity is equal to one then the cost of all routes is equal to the travel time to and from all patients. This is our baseline scenario and all other scenarios are compared to it.

When vehicles have capacity greater than one, we simplify the problem of patient transport to the deterministic Capacitated Vehicle Routing Problem (CVRP). The CVRP is a well known and studied problem in the vehicle routing literature. As we must solve medium to large CRVP instances thousands of times we do not make use of an industrial solver, such as Gurobi to solve to optimality, due to model runtime. Here we use a two-step heuristic approach. We first use Sequential Clarke-Wright Savings [12] and then use this as the initial ‘home base’ in a Iterated Local Search [13] meta-heuristic algorithm.

## 3 Results

### 3.1 Dialysis network

Currently the median travel time from home to dialysis unit (one way, with a single passenger) is 14 minutes. The minimum, lower quartile, upper quartile, and maximum travel times are 1, 9, 22, and 76 minutes.

Currently there is sufficient capacity for 668 dialysis patients in the outpatient sessions which are currently open, with 583 patients currently receiving dialysis (87% capacity utilisation).

Figures 2 to 4 show the effect of COVID progression if 80% of patients are infected over three months. If COVID progresses through 80% of the population in three months then, at the peak, there are up to about 125 COVID-positive patients (115-140 across the 30 model runs). Outpatients positives peak at about 65 (60-70) and inpatient positives peak at about 70 (60-85).

**Figure 2:**
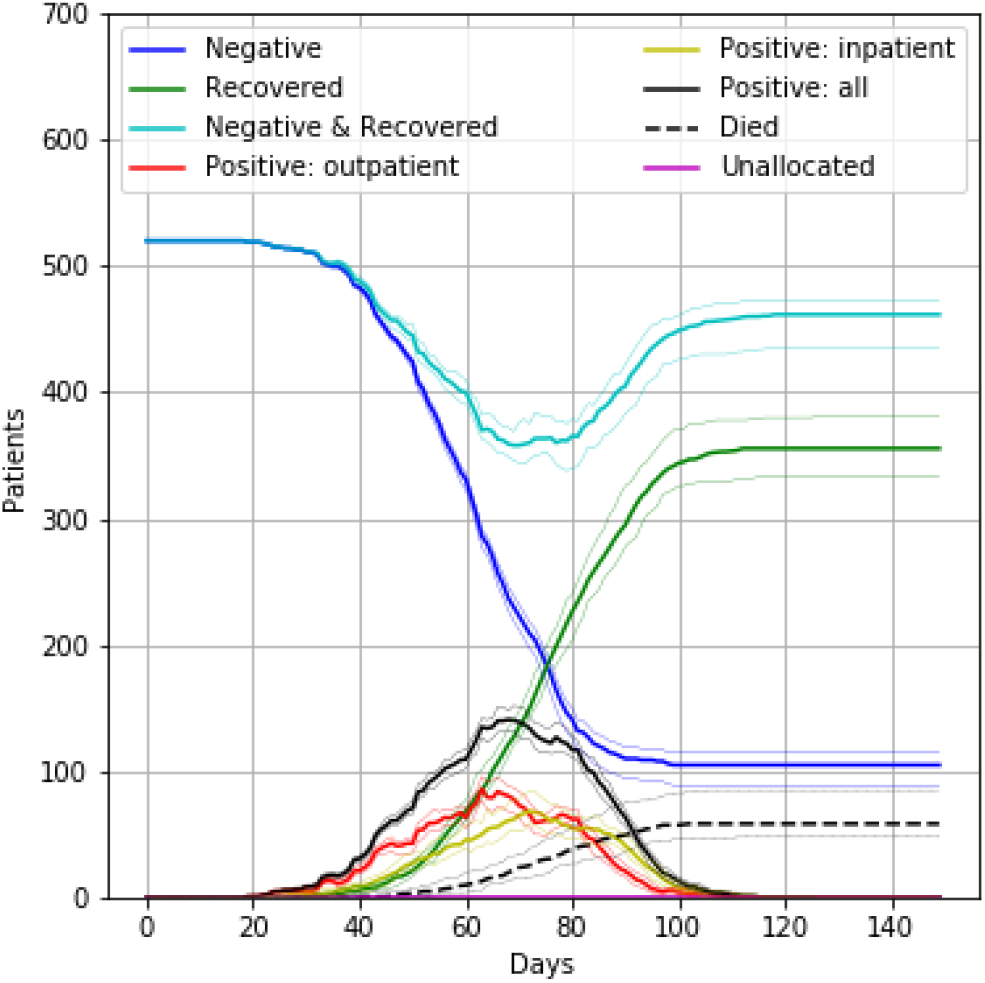
Progression of patient population through COVID infection, assuming 80% become infected over three months, with 15% mortality. The figure also shows the number of patients not allocated to a dialysis session at any time. The bold line shows the median results of 30 trials, and the fainter lines show the minimum and maximum from the 30 trials.

In the planned strategy of using half of one of the largest units (Queen Alexandra) for COVID-positive dialysis outpatients, and then using a second unit (Basingstoke, also providing up to half of its capacity for COVID-positive dialysis outpatient patients) for any excess, the dialysis system copes without any patients being unable to be allocated to a session (or without any need in dropping dialysis frequency). Workload in units that do not take COVID-positive outpatients will fall during the outbreak (though some work will flow back to them if they need to care for COVID-negative patients displaced from the units caring for COVID-positive patients).

One unit (Queen Alexandra) takes all COVID-positive inpatients in the model. The novel workload of treating COVID-positive patients who would otherwise not need inpatient care will likely stress inpatient care systems.

Outpatients may be displaced from their usual unit of care either because they need to travel to a COVID-positive session in another hospital, or because their unit has had to free up sessions for COVID-positive sessions. These patients typically require 20 minutes extra travel time to get to their temporary place of care (assuming they are travelling alone), with some requiring 50 minutes extra travel in each direction to/from dialysis.

### 3.2 Patient transport

Figures 5 and 6 illustrate the travel times distribution inbound, and inbound plus outbound (doubled inbound times), respectively, by transport capacity size of the COVID positive patient cohort.

**Figure 3:**
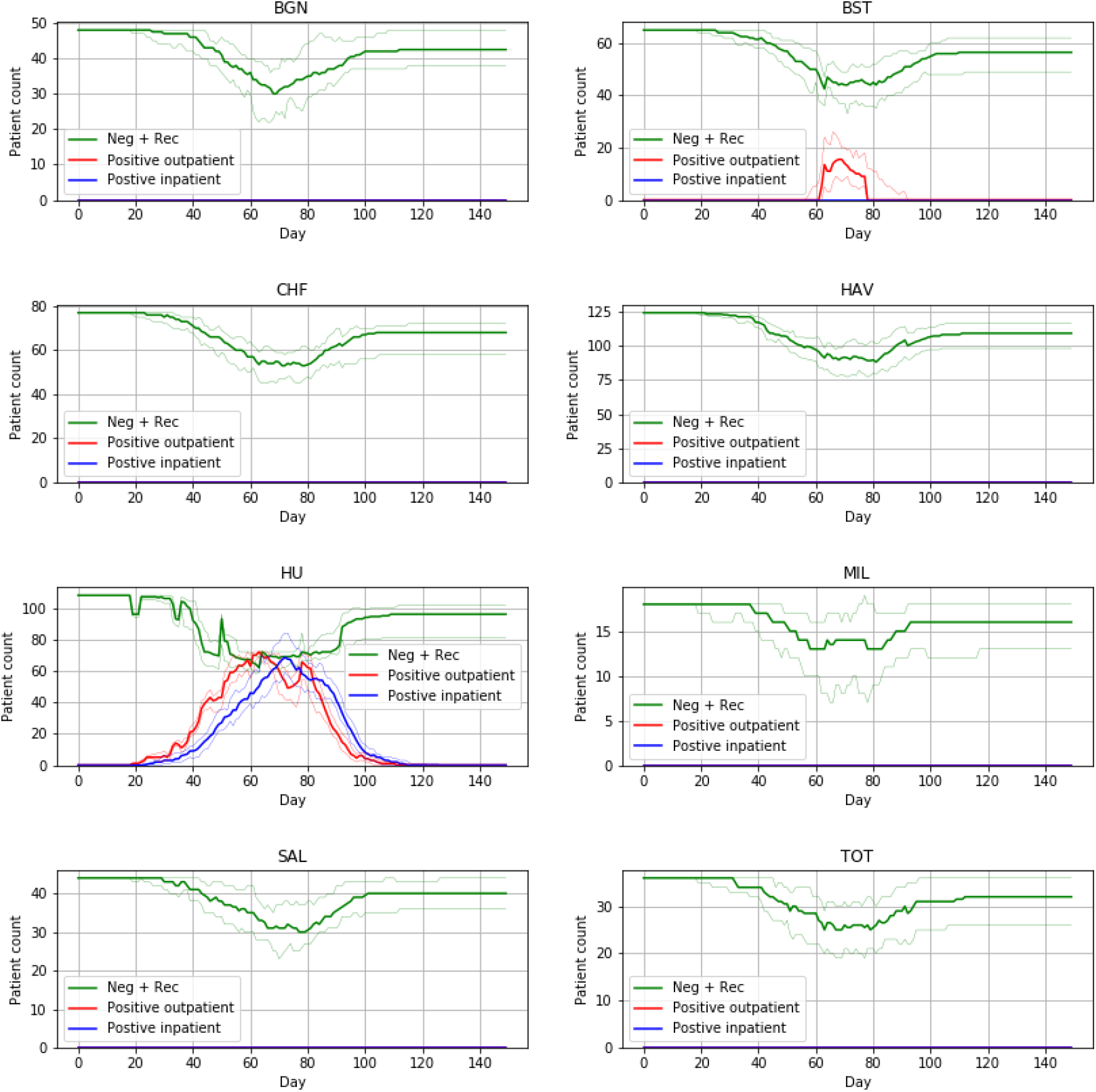
The number of patients (COVID negative, COVID-positive, COVID-inpatient, and COVID-recovered) allocated to each unit over time. The patient population progresses through infection over three months (with 80% infected). The bold line shows the median results of 30 trials, and the fainter lines show the minimum and maximum from the 30 trials.

**Figure 4:**
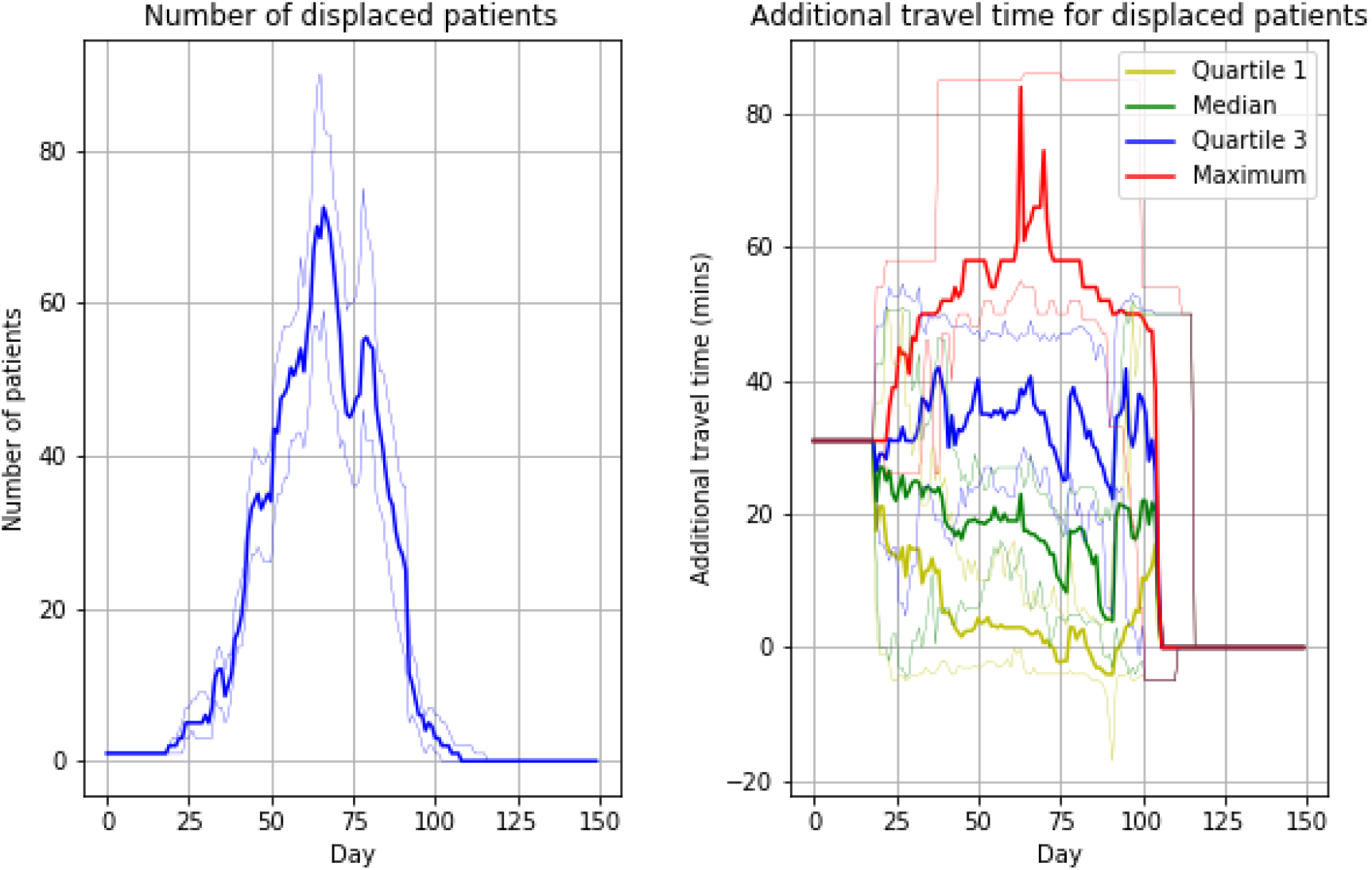
The number of patients displaced from their current unit (left panel) and the additional travel time to the unit of care (right panel) for displaced patients. These results do not include those receiving inpatient care. The patient population progresses through infection over three months (with 80% infected). The bold line shows the median results of 30 trials, and the fainter lines show the minimum and maximum from the 30 trials.

**Figure 5:**
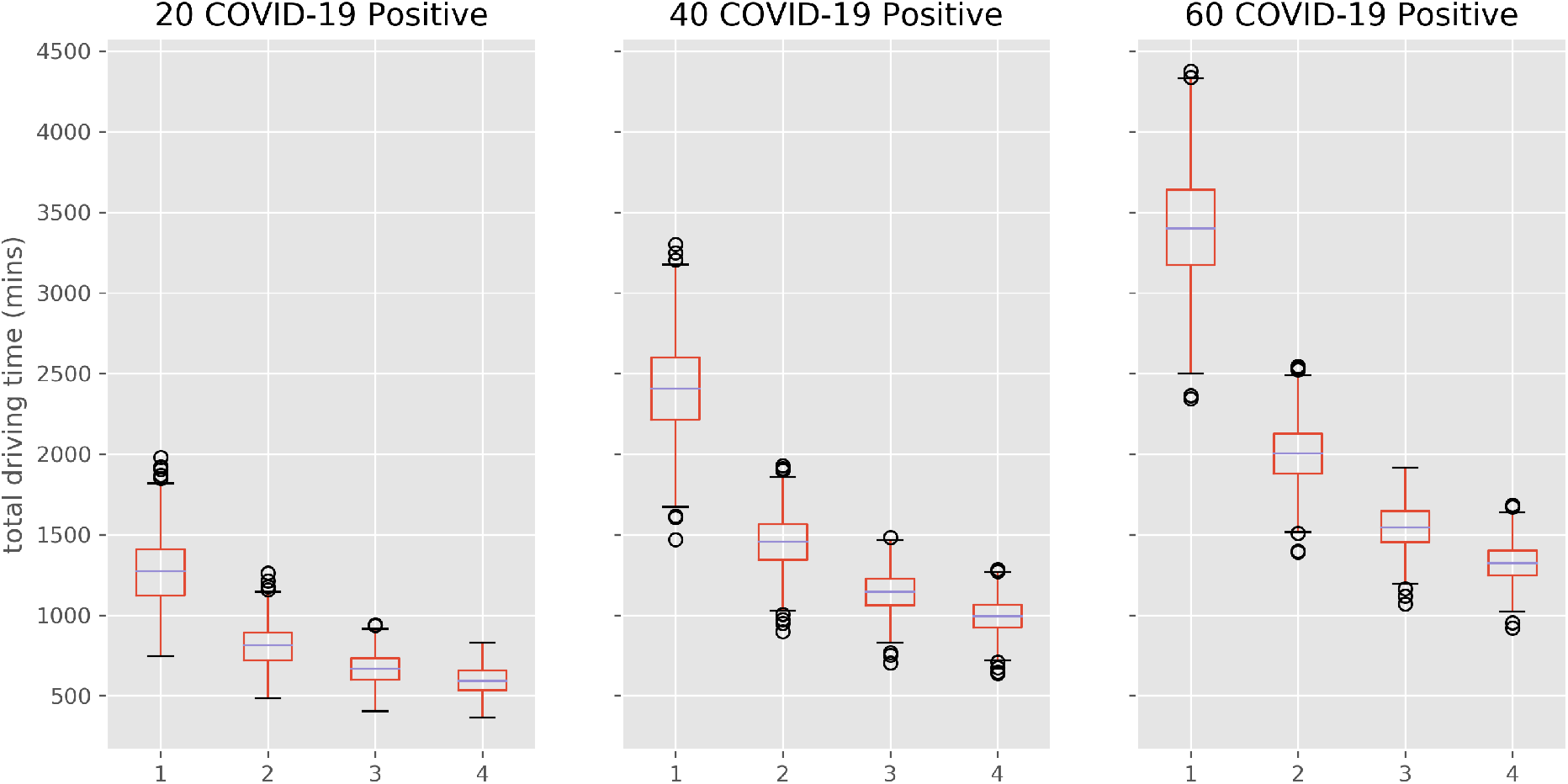
One-way ambulance transport time distributions (1000 model runs). Results compare population COVID +ive and ambulance seating capacity (e.g. 2 = 2 seats.) Figures do not include ambulance clean-down/turnaround time.

**Figure 6:**
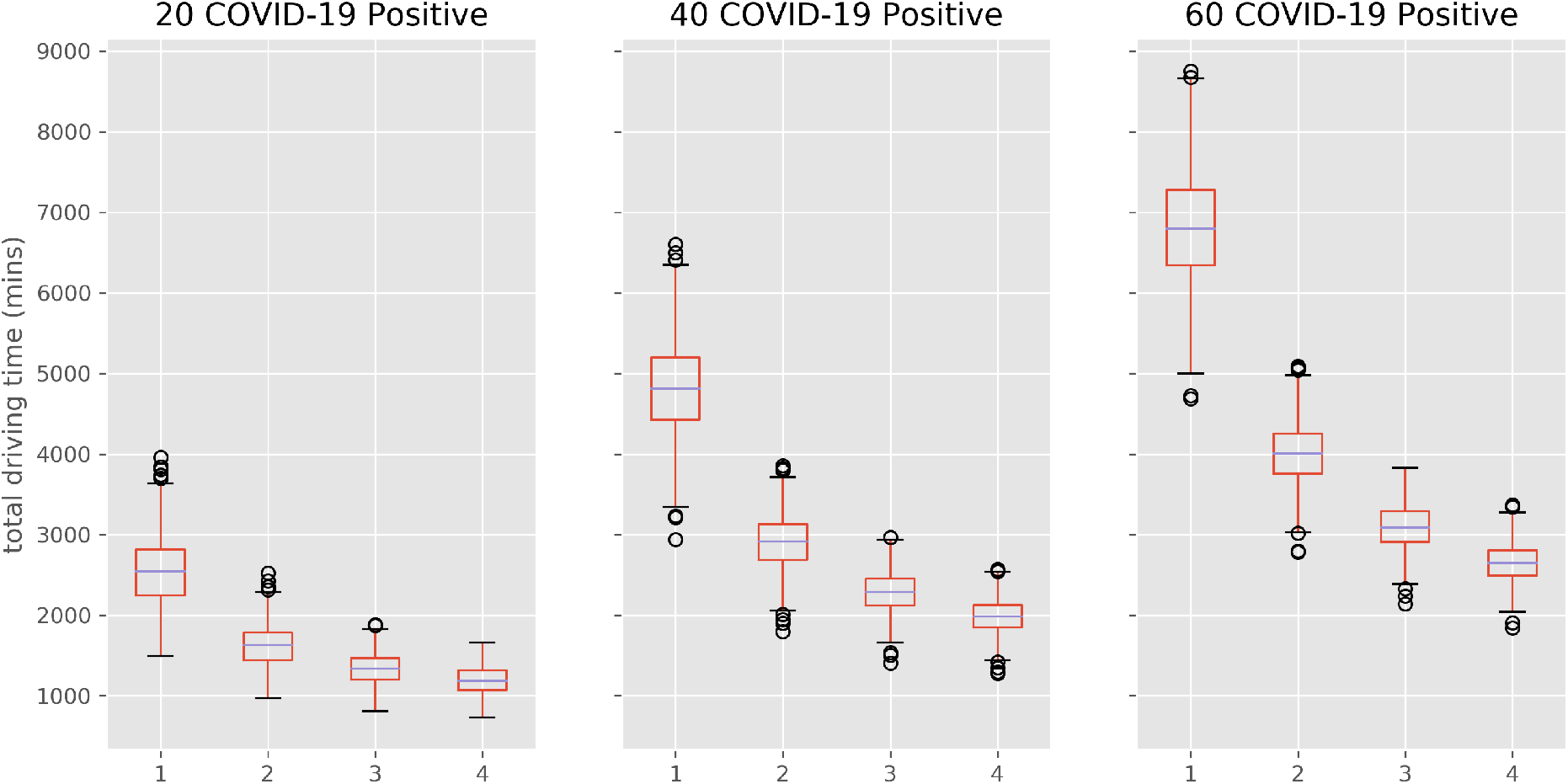
Two-way ambulance transport time distributions (1000 model runs). Results compare population COVID +ive and ambulance seating capacity (e.g. 2 = 2 seats.) Figures do not include ambulance clean-down/turnaround time.

A patient transport policy where a single patients are transported at a time has a median time substantially higher than all multi-occupancy policies. In all COVID caseload scenarios the largest improvement is seen when a additional patient is transported in each trip. Further improvement is seen if vehicle capacity is increased to three or four patients.

For example, if 40 COVID-positive patients need inbound and outbound transport, then a a median of 80.0 hours (inter-quartile range = 12.5 hours) of ambulance driving time (not including fixed drop-off and clean times) is required per day. If the transport capacity of vehicles is increased to two, three or four patient seating capacity, the median travel time requirements are reduced to 48 hours (a 40% reduction relative to single occupancy vehicles), 38 hours (52% reduction) and 33 hours (60% reduction), respectively.

## 4 Discussion

The results indicate that, if current outpatient capacity is maintained, the dialysis units should be able to cope with the worst-case scenario of rapid (three month) spread of COVID, but that workloads will shift to the central hospital. Coping with a rapid spread of COVID will require rapid reallocation of patients to different sessions and units, an effect likely to also impact on ambulance transfer services who will see journey times increase, and have reduced efficiency of having to split COVID positive and COVID-negative patients.

It appears likely that there will be significant inpatient pressures, with current capacity likely to be breached. It may be necessary to consider moving dialysis equipment during the peak COVID-positive workload when demand on units taking COVID-negative patients only will be reduced.

The current practice of transporting COVID-positive patients individually appears unsustainable. The results demonstrate that single seat ambulances face a challenge in transporting COVID-positive patients to and from the QA on a given day. In each scenario there is significant savings from using the additional ambulance capacity for more COVID-positive patients. In each scenario the biggest relative improvement is seen when capacity is increased to 2 seats (e.g. reducing ambulance drive time from 75 to about 45 hours per day for 40 patient two-way journeys). Increasing to 2-4seats has further benefit, but returns are diminishing.

### 4.1 Limitations of the study

A general limitation to these types of models is the level of uncertainty about the spread of COVID. We have therefore sought to model worst-case scenarios to enable contingency planning.

#### 4.1.1 Dialysis model

- The model assumes that patients can be re-allocated to units/sessions immediately. In practice changes to session allocation (e.g. shifting from COVID-negative to COVID-positive are likely to be made a little in advance.
- The results reported here assume that current capacity is maintained throughout the COVID outbreak. We have not modelled the effect of reductions in capacity that may be caused by staff shortages.
- We have not modelled timing of sessions, but the model progressively allocates COVID-positive sessions as needed, and we would assume that these sessions would come later in the day, enabling cleaning at the end of the day, ready for any COVID-negative session the next morning.
- We have not included home dialysis patients, which may affect inpatient demand. A likely worst-case scenario (with home dialysis patients following the transmission spread, and need for inpatient care, of the dialysis units, is that inpatient demand may be increased 15%.

#### 4.1.2 Transport model

- The findings provide estimates of total time patient transport ambulances will need to travel. They are not intended to provide recommendations of the minimum number of ambulances needed to maximise the number of appointments and/or shifts that run on time. A more accurate, but highly complex and time consuming, formulation of this problem is called the (static) dial-a-ride problem [21]. Dial-a-ride formulations explicitly take account of *time windows* for patient pickup and drop-off and *maximum patient ride-time*. Further dynamic complexity would be required in order to incorporate two-way patient journeys.
- The route optimisation uses two well known heuristics. A heuristic algorithm offers a fast method to obtain a good solution, but it does not guarantee an optimal solution i.e the shortest possible travel time achieved by optimal assignment of patients to routes. It is possible to solve the CRVP with 100 nodes to ‘optimality’ using industrial solvers such Gurobi. We chose a heuristic approach primarily for solution speed as we made no assumptions about the size of problem others networks could face internationally. We note that a possible improvement to the approach could be to switch to the Parallel version of Clarke-Wright savings.
- Figures for inward and outwards journeys do no explore the potential efficiencies of dropping patients back at their homes (after dialysis) and picking new patients up at the same time.

## Data Availability

All data and code are available and links/doi is available in the article.

https://git.exeter.ac.uk/tmwm201/dialysis-service-delivery-covid19

## Footnotes

### Contributions

All authors conceived the aims of the study. MA and TM conducted the analyses and wrote the paper. AB, SD and JW provided clinical and health system scrutiny of the work. All authors were involved in the development and review of the manuscript.

### Disclaimer

The views expressed in this publication are those of the author(s) and not necessarily those of the National Health Service, the NIHR, or the Department of Health and Social Care.

### Patient consent

Not required

### Ethics

The research team had no access to individual patient records. All analyses were conducted with aggregate counts of patients per UK postcode sector.

### Code and Data Availability

The code and data used in this study is available at https://git.exeter.ac.uk/tmwm201/dialysis-service-delivery-covid19.

## References

[1] N. M. Ferguson, D. Laydon, G. Nedjati-Gilani, et al. “Report 9. Impact of non-pharmaceutical interventions (NPIs) to reduce COVID-19 mortality and healthcare demand”. en. In: Imperial College Preprint (2020). url: https://www.imperial.ac.uk/media/imperial-college/medicine/sph/ ide/gida-fellowships/Imperial-College-COVID19-NPI-modelling-16-03-2020.pdf.

[2] N. I. for Health and C. Excellence. “COVID-19 rapid guideline: dialysis service delivery NICE guideline [NG160] Published date: 20 March 2020”. In: NICE (2020). url: https://www.nice.org.uk/guidance/ng160.

[3] C. Basile, C. Combe, F. Pizzarelli, et al. “Recommendations for the prevention, mitigation and containment of the emerging SARS-CoV-2 (COVID-19) pandemic in haemodialysis centres”. In: Nephrology Dialysis Transplantation (Mar. 2020). doi: 10.1093/ndt/gfaa069. url: https://doi.org/10.1093/ndt/gfaa069.

[4] A. S. Kliger and J. Silberzweig. “Mitigating Risk of COVID-19 in Dialysis Facilities”. In: Clinical Journal of the American Society of Nephrology (2020). doi: 10.2215/CJN.03340320. url: https://cjasn.asnjournals.org/content/early/2020/03/20/CJN.03340320.

[5] C. S. Currie, J. Fowler, K. Kotiadis, et al. “How simulation modelling can help reduce the impact of COVID-19”. In: Journal of Simulation (2020). doi: 10.1080/17477778.2020.1751570. url: https://doi.org/10.1080/17477778.2020.1751570.

[6] M. Pitt, T. Monks, S. Crowe, et al. “Systems modelling and simulation in health service design, delivery and decision making”. In: BMJ Quality & Safety 25.1 (2016), pp. 38–45. doi: 10.1136/bmjqs-2015-004430. url: https://qualitysafety.bmj.com/content/25/1/38.

[7] S. C. Brailsford, P. R. Harper, B. Patel, et al. “An analysis of the academic literature on simulation and modelling in health care”. In: Journal of Simulation 3.3 (2009), pp. 130–140. doi: 10.1057/jos.2009.10. url: https://doi.org/10.1057/jos.2009.10.

[8] M. M. Günal and M. Pidd. “Discrete event simulation for performance modelling in health care: a review of the literature”. In: Journal of Simulation 4.1 (2010), pp. 42–51. doi: 10.1057/jos.2009.25. url: https://doi.org/10.1057/jos.2009.25.

[9] S. Mohiuddin, J. Busby, J. Savović, et al. “Patient flow within UK emergency departments: a systematic review of the use of computer simulation modelling methods”. In: BMJ Open 7.5 (2017). doi: 10.1136/bmjopen-2016-015007. url: https://bmjopen.bmj.com/content/7/5/e015007.

[10] P. Roderick, R. Davies, C. Jones, et al. “Simulation model of renal replacement therapy: predicting future demand in England”. In: Nephrology Dialysis Transplantation 19.3 (Mar. 2004), pp. 692–701. doi: 10.1093/ndt/gfg591. url: https://doi.org/10.1093/ndt/gfg591.

[11] M. Allen, A. Spencer, A. Gibson, et al. “Right cot, right place, right time. Using neonatal care data and computer simulation to improve the design and organisation of neonatal care networks”. In: Health Serv Deliv Res (20 2015). doi: 10.3310/hsdr03200.

[12] G. Clarke and G. Wright. “Scheduling of vehicles from a central depot to a number of delivery points”. In: Operations Research 12 (1964), pp. 568–581.

[13] H. R. Lourenço, O. C. Martin, and T. Stützle. “Iterated Local Search”. In: Handbook of Metaheuristics. Ed. by F. Glover and G. A. Kochenberger. Boston, MA: Springer, 2003, pp. 320–353. doi: 10.1007/0-306-48056-5_11.

[14] M. Allen and T. Monks. COVID19 dialysis service delivery simulation models. Version v1.0.0. Apr. 2020. doi: 10.5281/zenodo.3760626. url: https://doi.org/10.5281/zenodo.3760626.

[15] Team SimPy. SimPy 3.0.11. https://simpy.readthedocs.io/en/latest/index.html. 2020.

[16] W. McKinney. “pandas: a foundational Python library for data analysis and statistics”. In: Python for High Performance and Scientific Computing 14 (2011).

[17] S. van der Walt, S. C. Colbert, and G. Varoquaux. “The NumPy Array: A Structure for Efficient Numerical Computation”. In: Computing in Science Engineering 13.2 (2011), pp. 22–30. doi: 10.1109/MCSE.2011.37.

[18] J. D. Hunter. “Matplotlib: A 2D graphics environment”. In: Computing in Science & Engineering 9.3 (2007), pp. 90–95. doi: 10.1109/MCSE.2007.55.

[19] T. Monks, C. S. M. Currie, B. S. Onggo, et al. “Strengthening the reporting of empirical simulation studies: Introducing the STRESS guidelines”. In: Journal of Simulation 13.1 (2019), pp. 55–67. doi: 10.1080/17477778.2018.1442155. url: https://doi.org/10.1080/17477778.2018.1442155.

[20] R. G. Sargent. “Verification and validation of simulation models”. In: Journal of Simulation 7.1 (2013), pp. 12–24. doi: 10.1057/jos.2012.20. url: https://doi.org/10.1057/jos.2012.20.

[21] J.-F. Cordeau and G. Laporte. “The dial-a-ride problem: models and algorithms”. In: Annals of Operations Research (1 2007), pp. 29–46. doi: 10.1007/s10479-007-0170-8. url: https://doi.org/10.1007/s10479-007-0170-8.

